# Claims-based reconstruction of the pancreatic cancer care pathway links centralization of care to survival

**DOI:** 10.64898/2026.10.17.26365423

**Authors:** Daniel Laubr, Aleš Tichopád, Marian Rybář, Zuzana Bielčiková, Martin Loveček, Martin Rožánek, Gleb Donin

## Abstract

**Background:** The organization of pancreatic cancer care may be associated with survival. Using health administrative claims data, we reconstructed the pathway from diagnosis to first-line therapy and examined associations of multidisciplinary team review and treatment at specialized centers with overall survival.

**Methods:** Longitudinal historical cohort study using anonymized health administrative claims data from six Czech health insurance funds, covering approximately 17% of patients diagnosed with pancreatic cancer nationally between 2018 and 2020. Adults with an incident ICD-10 C25 record confirmed by imaging and histopathology were followed from the index date (the date of the first qualifying imaging or histopathological examination) until death, censoring, or 31 December 2021. Overall survival was analyzed using Kaplan–Meier methods and Cox regression adjusted for age and first-line therapy type.

**Results:** Of 1,255 patients, 920 (73.3%) received first-line therapy and 266 (21.2%) died untreated. Among treated patients, 436 (47.4%) underwent multidisciplinary team review, 335 (36.4%) underwent surgery, and 667 (72.5%) were treated at a complex oncological center. Surgery was associated with longer overall survival than chemotherapy (age-adjusted hazard ratio [HR] 0.32, P < 0.001). Treatment at a complex oncological center was associated with longer overall survival (adjusted HR 0.77, 95% confidence interval [CI] 0.64–0.92, P = 0.004; crude median 532 versus 394 days). Multidisciplinary team review was not significantly associated with overall survival (adjusted HR 0.86, 95% CI 0.73–1.01, P = 0.073).

**Conclusions:** Treatment at a complex oncological center was associated with longer survival, while fewer than half of patients had a recorded multidisciplinary team review. How care is organized, not only which therapy is delivered, is associated with outcome. The observational design and absence of key clinical covariates limit causal interpretation.

## 1 Introduction

Pancreatic cancer poses a formidable challenge in oncology due to its aggressive nature and limited treatment options, necessitating a comprehensive and multifaceted approach to improve patient outcomes (1, 2). As in other countries with growing life expectancy, the incidence of pancreatic cancer has increased by almost 50% since 2000 in the Czech Republic, with 7,559 new cases reported over a 3-year period between 2018 and 2020 (3).

Generally, in all developed healthcare systems, the clinical pathways of patients with pancreatic cancer typically begin with the presentation of symptoms, such as abdominal pain, weight loss, or jaundice, or as an incidental diagnosis. Upon seeking medical attention, the primary care physician may suspect pancreatic cancer and refer the patient to a gastroenterologist or an oncologist for further evaluation. The patient then undergoes a comprehensive diagnostic workup, including imaging tests such as mandatory computed tomography scans and ultrasonography, and sometimes other examinations such as magnetic resonance imaging and endoscopic biopsy.

According to local and international guidelines, once diagnosed, the patient should be reviewed by a multidisciplinary team (MDT), typically consisting of oncologists, surgeons, radiologists, and other specialists (4, 5, 6). MDT review is generally believed to benefit patients, in terms of superior survival compared with that of patients without MDT (7). However, it was also reported that MDT review and other clinical examinations may cause an unnecessary delay in patients where surgery offers an obvious benefit (8). Therefore, a question remains about whether MDT review constitutes a bottleneck for preselecting patients for surgery.

Similarly, it was repeatedly shown that centralization of pancreatic cancer treatment improves the outcomes of patients due to greater utilization of neoadjuvant therapy (9), changes in surgical patterns (10), abundant experience of resections (10, 11), and more frequent use of chemotherapy in patients with advanced pancreatic cancer (12).

The approaches of MDT review and centralized treatment are based on the comprehensive experience of large numbers of patients across various medical perspectives. Therefore, we consider both factors to be key components of strict observance of treatment guidelines.

Surgery is seen as the most favorable treatment for pancreatic cancer, with median overall survival (OS) times of 14.8 and 18.8 months in two recent meta-analyses (13, 14). The 1-year OS rate of patients undergoing radical resection is 52%–83%, compared to 29.9% for patients without surgical resection (14, 15). Adjuvant chemotherapy also significantly prolongs OS, with median values ranging from 28 to 54 months, depending on the type of chemotherapy used (16, 17). However, only 15%–20% of cases of pancreatic cancer are considered resectable (18). In pivotal trials of systemic therapy for metastatic pancreatic cancer, median OS ranged from 6.7 to 11.1 months (19, 20).

Health administrative claims (HAC) data, which are available in broad, standardized, and digital form, are now acknowledged as an unprecedented source of real-world evidence (21). Studies have proven the capacity of HAC data to deliver vital insights over time (22). The identification of critical points in the patients’ pathways could help to optimally manage large numbers of patients (23).

In this study, we examined the temporal and sequential aspects of care from the diagnosis of pancreatic cancer to first-line therapy (FLT), and the associations of MDT review and centralization of care with OS in routine clinical practice.

## 2 Materials and methods

### 2.1 Study design and data source

This was a retrospective longitudinal cohort study using real-world data. The study period ranged from January 2017 to December 2021. We utilized anonymized HAC data, comprising de-identified information about patient interactions with the healthcare system, from six health insurance companies in the Czech Republic. Patient data included age, sex, medical specialty, International Classification of Diseases, 10th revision (ICD-10) diagnoses, and the date of death where reported. The study is reported in accordance with the STROBE statement for observational studies (Supplementary Material 2).

#### 2.1.1 Ethics

This study was performed in line with the principles of the Declaration of Helsinki. The study was conducted retrospectively on fully anonymized health administrative claims data that contained no directly or indirectly identifying information, and for which no additional patient contact or intervention took place. The Ethics Committee of the Faculty of Biomedical Engineering, Czech Technical University in Prague, waived the requirement for ethical approval for this work under its statement dated 18 November 2024. The statement covers retrospective, non-interventional research using anonymized health administrative data conducted at the Department of Biomedical Technology under project CZ.02.01.01/00/23_025/0008743. The present study falls within this scope and complies with the conditions specified in the statement.

### 2.2 Patient selection

Complete HAC data were defined as at least one record for each calendar year between the year prior to the event date and the year of the censoring date.

In order to address the risk of misclassification, we included patients with at least one inpatient and/or two outpatient records (documents) with the ICD-10 code C25 separated by a minimum interval of 30 days.

We omitted the year 2017 to ensure that only incident patients were included (i.e., a washout window of a minimum of 365 days), and the year 2021 to allow evaluation of patient outcomes. Patients were excluded if they did not have complete HAC data or had inconsistent pancreatic cancer diagnosis codes.

Imaging and histopathology within 60 days with a C25 diagnosis had to be reported on those examinations or on any other claim document within 30 days following the later of the two.

Death was the event of interest. Patients without a recorded death were censored on the last day of the last calendar year with available patient data or on 31 December 2021, whichever came first. This date was established as the censoring date.

The study design and cohort selection are depicted in Figure 1, as suggested by Schneeweiss et al. (24). Cohort entry was defined as the first occurrence of pancreatic cancer-related imaging and histopathological examinations within the selected period. The index date was defined as the date of the first of these examinations. Patients were excluded from the study if they had events prior to January 2018, were <18 years old at the index date, or had records of pancreatic cancer-related surgery, chemotherapy, or radiotherapy in the year prior to the index date.

**Figure 1.**
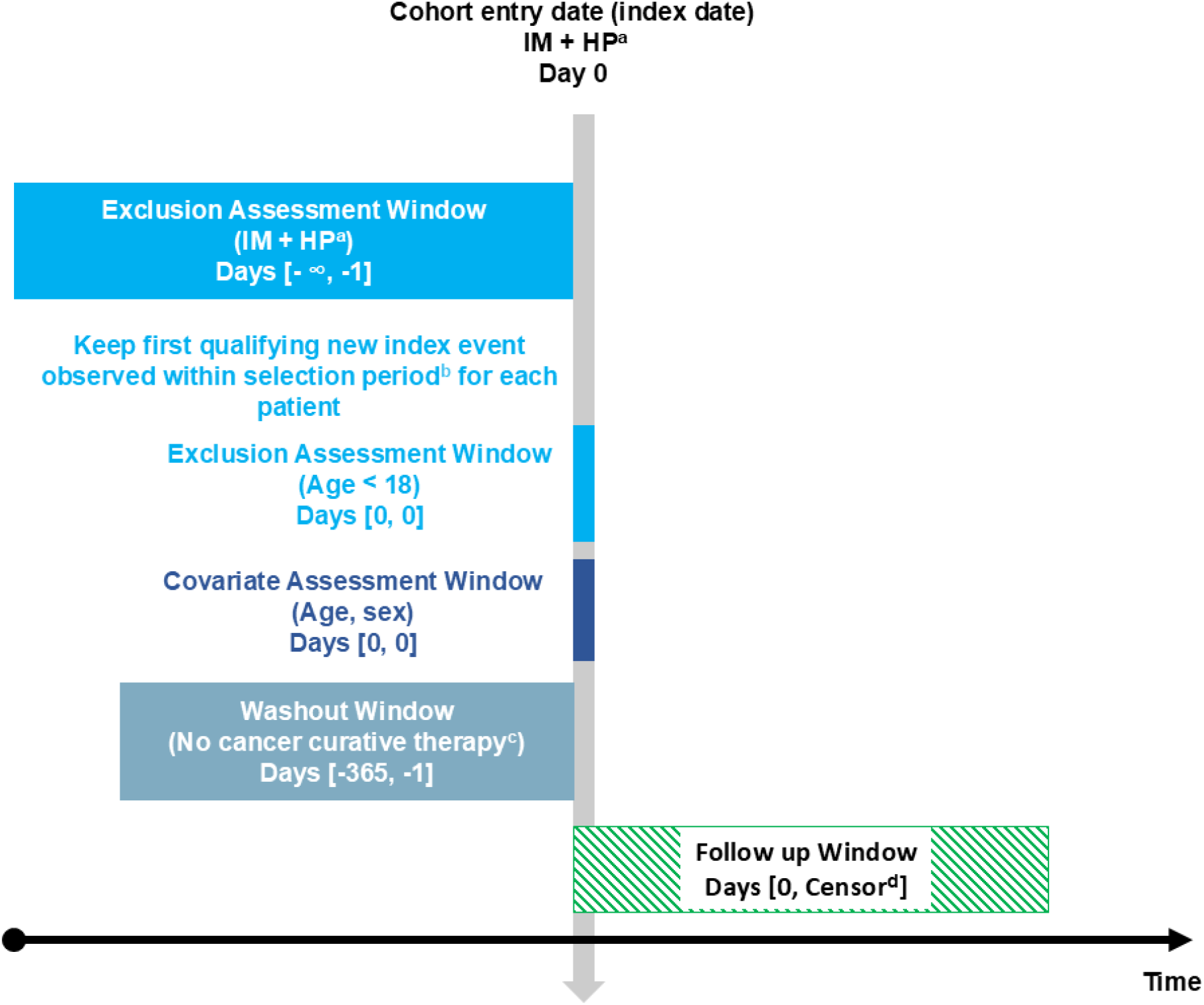
Overview of study design and cohort selection. (a) Imaging and histopathological examinations accompanied by ICD-10 code C25 within the specified time window. (b) January 2018 to December 2020. (c) Any chemotherapy, radiotherapy, or pancreatic cancer-related surgery. (d) Follow-up ended at death or censoring; patients without a recorded death were censored on the last day of the last calendar year with available data or 31 December 2021, whichever came first. HP, histopathological examination; ICD-10, International Classification of Diseases, 10th revision; IM, imaging examination.

### 2.3 Study outcomes

The primary objective was to assess the associations of MDT review and receipt of FLT at specialized centers, referred to as complex oncological centers (COCs) in the Czech Republic, with OS. A Cox proportional hazards model was utilized to analyze OS following MDT review and treatment in a COC. Age and the FLT (surgery, chemotherapy, or radiotherapy) were considered covariates. Patients were followed up from the index date until the occurrence of the event or censoring, as defined in Section 2.2.

The constructed clinical event sequences were analyzed as secondary objectives, in order to determine the baseline characteristics, the relative and absolute number of patients with different sequences of events, and the time between clinical events. For treated patients, we determined the actual treatment modality used as FLT, the time to treatment, and the percentage of patients who underwent MDT review. The time to treatment was defined as the time between the index date and the date of FLT.

Overall survival was evaluated using the Kaplan–Meier method. We performed survival analysis with stratification by the patient pathway, the presence of MDT review, the type of hospital that provided the FLT, and the type of FLT.

### 2.4 Statistical analysis

The initial data processing was performed in SAS (v9.4), and the statistical analyses were conducted using the R statistical language (version 4.3.1) for Windows (25). Variables with P < 0.25 in univariate analyses were entered into a multivariable Cox proportional hazards model together with the variables of interest with P < 0.05, to determine the adjusted associations of variables with outcome. The results of the multivariable Cox proportional hazards model are expressed as hazard ratios (HR) with 95% confidence intervals (CI) and P-values. Kaplan–Meier survival curves were plotted to graphically depict the variations in OS. Categorical variables are presented as frequencies and percentages. Continuous variables are presented as the mean and standard deviation or the median and range. Time differences are presented as the median and interquartile range. Median survival times are presented with 95% confidence intervals.

### 2.5 Use of generative artificial intelligence

A large language model ChatGPT (OpenAI; https://chatgpt.com; model/version: GPT-5.5) was used for formatting and language editing of the manuscript prior to submission. It was not used to generate, analyze, or interpret data, or to produce scientific content. The authors reviewed and edited all output and take full responsibility for the content of the article.

## 3 Results

### 3.1 Patients

A total of 2,426 patients with a first record of the ICD-10 code C25 between 2018 and 2020 were identified and retrieved from the database (Figure 2). Of these, 908 (37.4%) had no record of pancreatic cancer-related imaging and/or histopathological examination within the selection period, 212 (8.7%) had undergone curative cancer treatment before the index date, and 51 (2.1%) had the ICD-10 code C25 recorded before the index date. Thus, 1,255 (51.7%) patients satisfied the study’s inclusion criteria and comprised the study cohort. The median age of the study cohort was 68 years (interquartile range [IQR] 60–73 years), 631 (50.3%) were women, and 624 (49.7%) were men.

**Figure 2.**
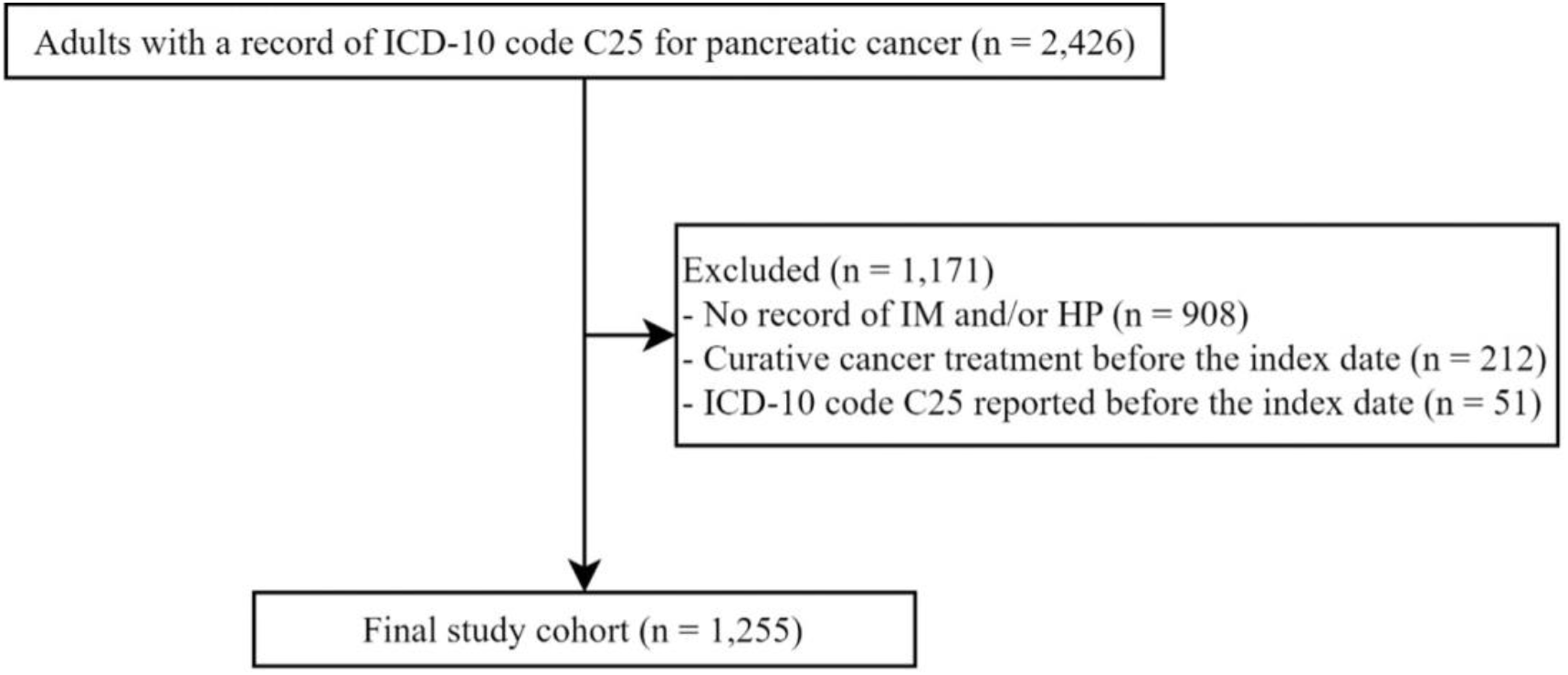
Patient disposition and eligibility. HP, histopathological examination; ICD-10, International Classification of Diseases, 10th revision; IM, imaging examination.

### 3.2 Patient pathways

Figure 3 depicts the generalized pancreatic cancer patient pathways prior to or during initial management. Out of 1,255 patients with pancreatic cancer, less than half (46.8%) were evaluated by MDT review and 38.6% received treatment without a recorded MDT review; 73.3% underwent FLT and 21.2% of patients died without receiving any treatment. Furthermore, 5.5% of patients were censored (Figure 3A).

**Figure 3.**
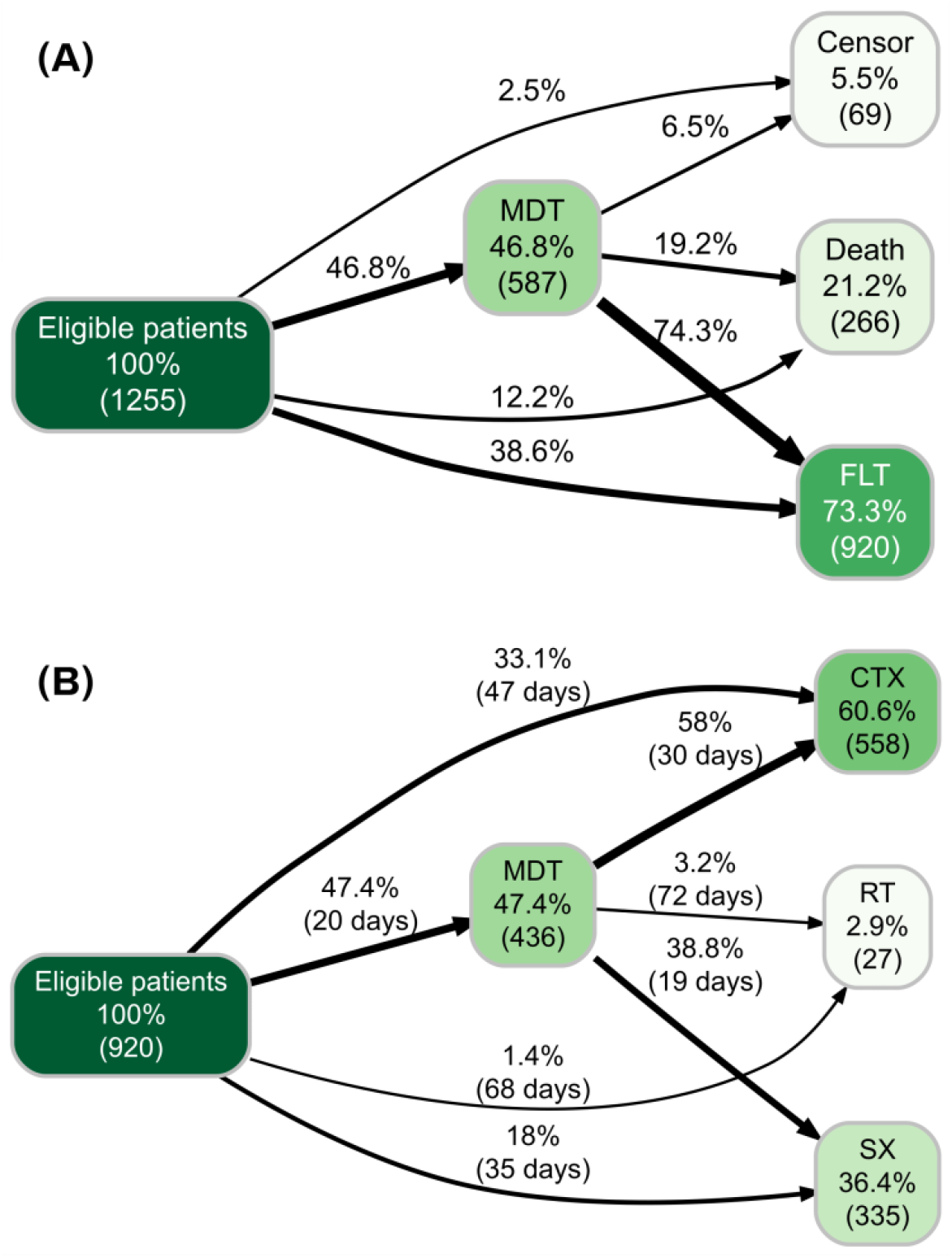
Pancreatic cancer care pathways from the index date to first-line therapy. (A) Whole study cohort. (B) Patients who received FLT. Node values show patient counts and percentages. Percentages on connecting lines use the preceding node as the denominator. Time intervals shown on connecting lines are median numbers of days between events. Medians of individual intervals were estimated separately and do not sum to the median total time from the index date to treatment. CTX, chemotherapy; FLT, first-line therapy; MDT, multidisciplinary team; RT, radiotherapy; SX, surgery.

Of the 920 treated patients, 60.6% underwent chemotherapy, 36.4% underwent surgery, and 2.9% underwent radiotherapy as FLT (Figure 3B). MDT review was performed in 47.4% of treated patients, increasing from 45.0% in 2018 to 49.5% in 2020 (Figure S1, Supplementary Material 1). A total of 72.5% (N = 667) of the treated patients underwent FLT at a COC.

Among the 436 treated patients whose care was based on an MDT decision, 169 (38.8%) underwent primary surgery, 253 (58.0%) underwent chemotherapy, and 14 (3.2%) underwent radiotherapy. Among the 484 treated patients without MDT review, the corresponding numbers were 166 (34.3%) for surgery, 305 (63.0%) for chemotherapy, and 13 (2.7%) for radiotherapy. Expressed relative to each treatment modality, MDT review preceded 50.4% of all surgeries, 45.3% of all courses of chemotherapy, and 51.9% of all courses of radiotherapy.

### 3.3 Time to treatment

In patients treated without MDT review, the median time from the index date to first-line therapy was 35 days for surgery, 47 days for chemotherapy, and 68 days for radiotherapy. In patients with MDT review, the corresponding median times were 44, 56, and 102 days, respectively. Among treated patients with MDT review (n = 436), the median time from the index date to MDT review was 20 days, and the median time from MDT review to first-line therapy was 19 days for surgery, 30 days for chemotherapy, and 72 days for radiotherapy (Figure 3B).

### 3.4 Patient prognosis

Among the 1,255 analyzed patients, one-fifth (21.2%) died without any treatment. The survival data of the 920 treated patients are summarized in Figure 4 according to the type of FLT, and are presented in more detail in Figure S2 and Table S2 (Supplementary Material 1).

**Figure 4.**
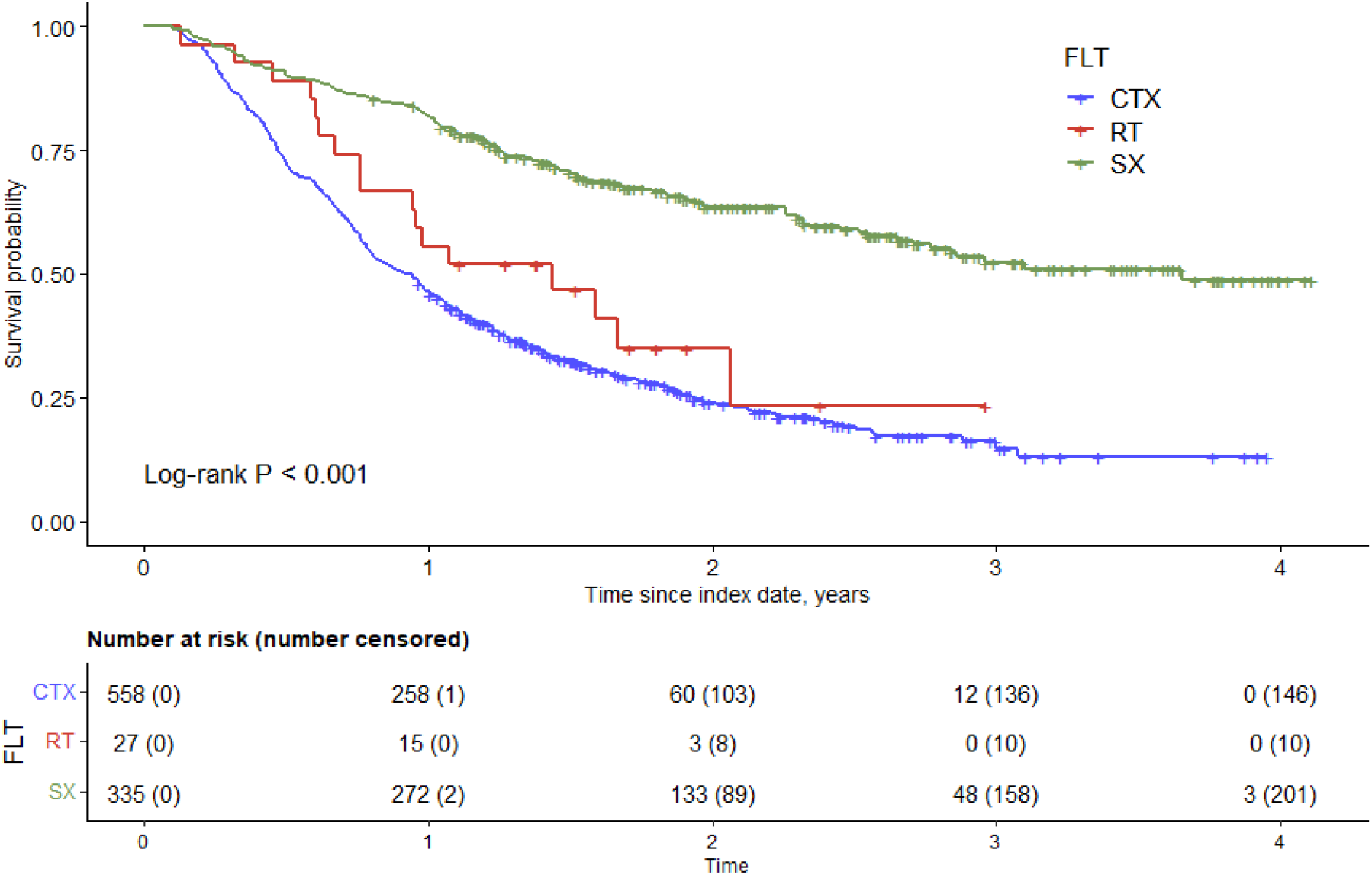
Kaplan–Meier estimates of overall survival in treated patients by type of first-line therapy. CTX, chemotherapy; FLT, first-line therapy; RT, radiotherapy; SX, surgery.

Surgical treatment was associated with better OS compared with chemotherapy, with an age-adjusted HR of 0.32 (P < 0.001). Among patients who underwent surgery, the median survival time was 1,333 days (95% CI 1,037–not reached [nr]) in those with MDT review versus 1,130 days (95% CI 856–nr) in those without MDT review. The median survival time of patients who underwent chemotherapy without surgery was 338 days (95% CI 293–374).

MDT review was not significantly associated with OS after adjustment for age and treatment type (HR 0.86, 95% CI 0.73–1.01, P = 0.073); crude median survival times were 531 versus 443 days in patients with and without MDT review. Treatment at a COC was associated with longer OS after adjustment for age and treatment type (HR 0.77, 95% CI 0.64–0.92, P = 0.004); crude median survival times were 532 versus 394 days in patients treated at COCs and non-COCs, respectively. Unadjusted Kaplan–Meier survival curves stratified by MDT review and treatment at a COC are presented in Figure S4 (Supplementary Material 1). Survival estimates by treatment type are shown in Table 1.

**Table 1.** Association of multidisciplinary team review and centralization of care with survival in treated patients.

|  | **MDT vs non-MDT** |  | **COC vs non-COC** |  |
| --- | --- | --- | --- | --- |
|  | **MDT** | **Non-MDT** | **COC** | **Non-COC** |
| Adjusted HR (95% CI)<sup>a</sup> | 0.86 (0.73–1.01) (*P* = 0.073) |  | 0.77 (0.64–0.92) (*P* = 0.004) |  |
| Crude median survival time, days (95% CI) | 531 (461–688) | 443 (385–532) | 532 (466–633) | 394 (301–462) |
| **Adjusted median survival time, days (95% CI)** |  |  |  |  |
| CTX | 374 (334–428) | 331 (286–367) | 368 (344–409) | 287 (262–352) |
| SX | nr (1095–nr) | 1130 (914–nr) | nr (1095–nr) | 987 (757–nr) |
| RT | 522 (358–1333) | 446 (307–1011) | 494 (352–1123) | 383 (274–857) |

<sup>a</sup> a Cox proportional hazards regression adjusted for age and treatment type. CI, confidence interval; COC, complex oncological center; CTX, chemotherapy; HR, hazard ratio; MDT, multidisciplinary team; nr, not reached; RT, radiotherapy; SX, surgery.

## 4 Discussion

In the midst of the ongoing and intense discussions about the optimal utilization of limited resources in healthcare, particularly addressing the shortage in specialists and hospital capacities, insights into the spatial and temporal aspects and outcomes of patient pathways are becoming increasingly valuable. The objective of our work was to obtain real-world evidence regarding the outcomes of patients with pancreatic cancer in relation to their healthcare pathways from the index date to FLT. In particular, we investigated the effects of the presence of MDT review and centralized care on patient survival. We also sought to compare the outcomes of patients who underwent surgery with those who did not.

The Czech Republic is among the top three countries with the highest incidence and mortality for pancreatic cancer (26). Our study utilized data from approximately 17% of patients diagnosed with pancreatic cancer between 2018 and 2020. The median age of 68 years in our cohort is consistent with reported median ages ranging from 57–74 years across various European patient populations (27).

Over 21% of patients died without receiving any treatment. This aligns with data published by King et al. (28), who reported that 26% of patients are not eligible for treatment for various reasons. Because the HAC dataset lacked information about the performance status of patients, their comorbidities, or preferences, we were unable to determine the specific reasons why these patients were not treated.

The National Comprehensive Cancer Network guidelines consider that MDT review is necessary, and the Chinese Medical Association recommends incorporating MDT review into the treatment flow for all patients with pancreatic cancer (5, 6). Here, we show that the clinical reality in the Czech Republic differs from the anticipated goal, because only 46.8% of all patients and 47.4% of treated patients underwent MDT review. Furthermore, the composition of MDTs varies, and in our opinion most MDTs in the Czech Republic lack nurses, psychologists, and other experts focused on the patient’s quality of life or the care of patients indicated for symptomatic treatment. Therefore, MDT review is probably not that common in patients with poor performance status. We found that 74.3% (N = 436) of patients discussed by an MDT were indicated for treatment. This is in line with the assumption that MDT review is often indicated for patients with a more favorable prognosis. However, the question arises as to why 484 patients (52.6% of all treated patients, and 38.6% of the whole cohort) were treated without being discussed by an MDT. Patients referred for treatment may differ in characteristics that were not captured in the claims data, irrespective of whether they underwent MDT review. Although we found an increase in the percentage of patients discussed by an MDT between 2018 and 2020, the number of patients referred for treatment via MDT review should be considerably higher. According to the Netherlands Cancer Registry, 88.8% of patients treated with curative intent were discussed at an MDT (29). This is particularly relevant in the context of the number of operations performed.

Two prominent high-volume cancer facilities in Ireland reported a surgery rate of 49% for stage I–III patients since 2010 (30). By contrast, a lower rate of 15.8%, irrespective of disease stage, was reported in the aforementioned Dutch study (29). In our cohort, 36.4% of treated patients underwent surgery as FLT. These proportions are not directly comparable because the denominators and stage distributions differ. Survival estimates do not resolve the absence of information on disease stage and R0 resections.

We also analyzed whether MDT review was associated with the time to treatment. The total time to surgery was longer in patients with MDT review than in those without (44 vs. 35 days), and the median time (20 days) from the index date to MDT review remains contentious. One might assume that the waiting list is independent of MDT review. Therefore, if the decision to undergo surgery is made at MDT review, it raises the question of why the time from MDT review to surgery is much shorter (19 days) than the time between diagnosis and surgery in patients without MDT review (35 days). We assume that surgery is prioritized after MDT review to avoid further delay. Another possible explanation is that MDT review might be favored at larger centers. It is probable that specialized centers have greater facilities, allowing shorter waiting times than smaller centers with fewer surgeons. Alternatively, smaller hospitals without an MDT need to refer patients to centers with surgical facilities, prolonging the waiting time to surgery. These possible explanations were not directly tested in the present study.

Similar explanations may also apply to the larger capacity of specialized centers compared with smaller hospitals, and prioritizing patients from MDT review may explain the variability in waiting times among patients indicated for chemotherapy (56 vs. 47 days with and without MDT review). We know from daily practice that patients with a confirmed diagnosis may be asked to find an oncologist by themselves rather than being recommended to visit a particular doctor. This might prolong the waiting time enormously.

Unlike for surgery and chemotherapy, the technical capacity for performing radiotherapy exceeds the personnel capacity. This may explain why the waiting times for radiotherapy were the longest of all treatment modalities, both with and without MDT review (102 and 68 days, respectively). The difference between these two groups should be interpreted with caution, as it is based on only 14 and 13 patients.

Published results on the correlations between wait times and the prognosis of patients with pancreatic cancer are inconsistent, but they typically define the time to treatment as the time from symptom onset or diagnosis to treatment, irrespective of MDT review. Some studies suggest that initiating therapy within 6 weeks from diagnosis is associated with improved survival (31), but others indicated that the length of the diagnostic process does not influence disease prognosis (32, 33, 34). Numerous variables, including the presence of metastases and the nature and type of symptoms, may contribute to variations in the wait time and its clinical significance (35).

The prognosis of Czech patients is comparable with international data. The survival after surgery reached 3.1–3.7 years, but the survival of patients who underwent chemotherapy, without surgery, was approximately 1 year (Table S2, Supplementary Material 1). Given that our analysis exclusively focused on the patient’s pathway from diagnosis to FLT, the impact of adjuvant therapy on the OS of operated patients remains unclear in this study. Additionally, it is important to distinguish between neoadjuvant and palliative chemotherapy.

Several studies have highlighted the prognostic significance of centralized care (9, 10, 11, 12). While most European countries have faced challenges in centralizing pancreatic surgery to high-volume centers for various reasons (36), the Czech Republic established a network of COCs in 2005. Nevertheless, the centralization of care in COCs does not fully replicate the centralization of pancreatic surgery. In our data, longer survival in COCs was observed for patients treated with systemic chemotherapy and in those who underwent surgery (Figure S3 and Table S3, Supplementary Material 1). Possible explanations, suggested by previous studies, include the use of more effective chemotherapy regimens (9), a reduced time to the initiation of chemotherapy, and its broader indication for patients with poorer performance status, the elderly, or those with metastatic disease, relative to non-COC settings (12). Surgery performed in a COC is associated with a greater likelihood of earlier and more successful surgery, with fewer postoperative complications, and facilitates adjuvant chemotherapy (10, 11).

MDT review was not significantly associated with OS in the adjusted analysis, and the confidence interval included no association. Interpretation is limited by possible differences in case mix and referral patterns. MDT review is not restricted to COCs, and the two exposures may be correlated. The timing of MDT review and treatment may also matter. These factors warrant further investigation before attributing survival differences to either aspect of care.

MDT involvement may facilitate referral to specialized units and access to surgery (37). However, the present observational analysis cannot establish that MDT review or centralization improves survival for every patient or disease stage. Further research should evaluate referral mechanisms and the timing of care while accounting for clinical differences between patient groups.

### 4.1 Limitations of the study

Our study has some technical, procedural (Table S4, Supplementary Material 1), and statistical limitations. As mentioned earlier, we lacked information on performance status, comorbidities, and patient preferences. Additionally, we did not have any data regarding disease stage and histopathological type. Although all types of surgery performed were curative in intent, we lacked information on their radicality. Information on the type of chemotherapy was available but was not included in our analysis. For interpretation, we assumed that patients who underwent surgery were in good health with non-metastatic pancreatic cancer, and that their surgery was radical. Patients receiving chemotherapy may have had unresectable or metastatic disease, but treatment intent could not be established from the present analysis. Distinguishing between neoadjuvant, adjuvant, and palliative therapy requires data analysis beyond the scope of our focus on FLT. These assumptions were not directly verifiable. Given these limitations, it is possible that the patients’ prognosis could be influenced, for example, by differences in histopathological subtype, non-radical surgery, indication for neoadjuvant chemotherapy (13), or the efficacy of adjuvant chemotherapy (16), which was not specifically analyzed in our study. A further limitation is that assignment to MDT review and to a COC was not randomized, so residual confounding by indication cannot be excluded. Despite these limitations, HAC data are very useful for understanding patient pathways and clinical outcomes with regard to particular milestones, enabling quality comparisons at national levels.

## 5 Conclusion

We found that approximately half of patients in our cohort, including those indicated for surgery, are not discussed at MDT meetings. Centralization of first-line treatment at complex oncological centers was independently associated with longer overall survival, whereas MDT review showed only a non-significant trend. The overall prognosis of Czech patients nevertheless remains consistent with international data. These findings indicate that the organization of pancreatic cancer care, and not only the therapy selected, is associated with patient outcome, and that wider use of MDT review represents an actionable target for improvement.

## Data availability statement

The health administrative claims data analyzed in this study remain the property of the participating health insurance companies and cannot be shared publicly. Aggregated results are reported in the article and Supplementary Material 1. The analysis code supporting the findings is available from the corresponding author upon reasonable request. Further information and requests should be directed to the corresponding author.

## Author contributions

DL: Conceptualization, Methodology, Investigation, Formal analysis, Writing – original draft, Writing – review & editing. AT: Conceptualization, Methodology, Formal analysis, Project administration, Writing – original draft, Writing – review & editing. Marian Rybář: Conceptualization, Methodology, Writing – review & editing. ZB: Writing – original draft, Writing – review & editing. ML: Writing – review & editing. GD: Conceptualization, Methodology, Investigation, Formal analysis, Writing – original draft, Writing – review & editing. Martin Rožánek: Project administration, Writing – review & editing. DL, AT, GD, ZB, and ML contributed to data interpretation. All authors read, critically revised, and approved the final version of the manuscript and agree to be accountable for all aspects of the work.

## Acknowledgments

The data were kindly provided by the Health Insurance Bureau in Prague, Czech Republic. The authors thank Nicholas D. Smith (PhD) for editorial support. The authors used ChatGPT (OpenAI; model GPT-5.5; https://chatgpt.com) to assist with language editing, manuscript formatting, and checking citations and references. The tool was not used to analyze primary research data. The authors retain full responsibility for the accuracy of all statements and references, the interpretation of the findings, and the integrity and final content of the manuscript.

## Consent for publication

The manuscript contains no data or images from which an individual person could be identified.

## Supplementary material

Supplementary Material 1 (PDF) - Supplementary tables and figures: Table S1 Variables related to the patient pathway; Table S2 Median survival of treated pancreatic cancer patients according to the patient pathway (N = 920); Table S3 Adjusted survival of patients with pancreatic cancer according to the treatment strategy; Table S4 Study limitations; Figure S1 Time trend in MDT reporting for treated pancreatic cancer patients (N = 920); Figure S2 Survival curves of treated pancreatic cancer patients according to the patient pathway (N = 920); Figure S3 Survival curves of treated pancreatic cancer patients according to centralization of care and the type of therapy; Figure S4 Unadjusted survival curves of treated pancreatic cancer patients.

Supplementary Material 2 (PDF) - STROBE checklist for cohort studies.

## References

1. Kolbeinsson HM, Chandana S, Wright GP, Chung M. Pancreatic cancer: a review of current treatment and novel therapies. J Invest Surg (2023) 36:2129884. doi: 10.1080/08941939.2022.2129884

2. Regel I, Mayerle J, Mahajan UM. Current strategies and future perspectives for precision medicine in pancreatic cancer. Cancers (2020) 12:1024. doi: 10.3390/cancers12041024

3. Institute of Health Information and Statistics of the Czech Republic. Epidemiology of malignant tumours in the Czech Republic. Based on the National Health Information System (2022). Available at: https://www.svod.cz (accessed October 11, 2022).

4. Bielcikova Z, Ulrych J, Fryba V, Hrabak P, Zubal M, Krizova L, et al. Multimodal approach in the treatment of generalised malignancies of the gastrointestinal tract. Gastroent Hepatol (2022) 76:409–417. doi: 10.48095/ccgh2022409

5. Tempero MA, Malafa MP, Chiorean EG, Czito B, Scaife C, Narang AK, et al. Pancreatic adenocarcinoma, version 1.2019. J Natl Compr Canc Netw (2019) 17:202–210. doi: 10.6004/jnccn.2019.0014

6. Oncologist Branch of Chinese Medical Association, Expert Committee on Pancreatic Diseases of China Association for the Promotion of International Health Care Exchange, Abdominal Cancer Expert Committee of China Medical Education Association. Expert consensus on the MDT model of pancreatic cancer in China (2020 edition). Zhonghua Zhong Liu Za Zhi (2020) 42:531–536 (in Chinese). doi: 10.3760/cma.j.cn112152-20200310-00192

7. Katz MHG, Wang H, Fleming JB, Sun CC, Hwang RF, Wolff RA, et al. Long-term survival after multidisciplinary management of resected pancreatic adenocarcinoma. Ann Surg Oncol (2009) 16:836–847. doi: 10.1245/s10434-008-0295-2

8. Cooper M, Newman NA, Ibrahim AM, Lam E, Herman JM, Singh VK, et al. Unnecessary tests and procedures in patients presenting with solid tumors of the pancreas. J Gastrointest Surg (2013) 17:1218–1223. doi: 10.1007/s11605-013-2213-6

9. Hsu DS, Kumar NS, L. ST, Chang AL, Kazantsev G, Spitzer AL, et al. Centralization of pancreatic cancer treatment within an integrated healthcare system improves overall survival. Am J Surg (2022) 223:1035–1039. doi: 10.1016/j.amjsurg.2021.09.036

10. Lemmens VE, Bosscha K, van der Schelling G, Brenninkmeijer S, Coebergh JW, de Hingh IH. Improving outcome for patients with pancreatic cancer through centralization. Br J Surg (2011) 98:1455–1462. doi: 10.1002/bjs.7581

11. de Wilde RF, Besselink MG, van der Tweel I, de Hingh IH, van Eijck CH, Dejong CH, et al. Impact of nationwide centralization of pancreaticoduodenectomy on hospital mortality. Br J Surg (2012) 99:404–410. doi: 10.1002/bjs.8664

12. Faluyi O, Connor J, Chatterjee M, Ikin C, Wong H, Palmer DH. Advanced pancreatic adenocarcinoma outcomes with transition from devolved to centralised care in a regional cancer centre. Br J Cancer (2017) 116:424–431. doi: 10.1038/bjc.2016.406

13. Versteijne E, Vogel JA, Besselink MG, Busch ORC, Wilmink JW, Daams JG, et al. Meta-analysis comparing upfront surgery with neoadjuvant treatment in patients with resectable or borderline resectable pancreatic cancer. Br J Surg (2018) 105:946–958. doi: 10.1002/bjs.10870

14. Pecorelli N, Licinio AW, Guarneri G, Aleotti F, Crippa S, Reni M, et al. Prognosis of upfront surgery for pancreatic cancer: a systematic review and meta-analysis of prospective studies. Front Oncol (2022) 11:812102. doi: 10.3389/fonc.2021.812102

15. Nymo LS, Myklebust TÅ, Hamre H, Møller B, Lassen K. Treatment and survival of patients with pancreatic ductal adenocarcinoma: 15-year national cohort. BJS Open (2022) 6:zrac004. doi: 10.1093/bjsopen/zrac004

16. Niesen W, Hank T, Büchler M, Strobel O. Local radicality and survival outcome of pancreatic cancer surgery. Ann Gastroenterol Surg (2019) 3:464–475. doi: 10.1002/ags3.12273

17. Conroy T, Hammel P, Hebbar M, Ben Abdelghani M, Wei AC, Raoul JL, et al. FOLFIRINOX or gemcitabine as adjuvant therapy for pancreatic cancer. N Engl J Med (2018) 379:2395–2406. doi: 10.1056/NEJMoa1809775

18. van Veldhuisen E, van den Oord C, Brada LJ, Walma MS, Vogel JA, Wilmink JW, et al. Locally advanced pancreatic cancer: work-up, staging, and local intervention strategies. Cancers (2019) 11:976. doi: 10.3390/cancers11070976

19. Conroy T, Desseigne F, Ychou M, Bouché O, Guimbaud R, Bécouarn Y, et al. FOLFIRINOX versus gemcitabine for metastatic pancreatic cancer. N Engl J Med. (2011) 364:1817–1825. doi: 10.1056/NEJMoa1011923

20. Von Hoff DD, Ervin T, Arena FP, Chiorean EG, Infante J, Moore M, et al. Increased survival in pancreatic cancer with nab-paclitaxel plus gemcitabine. N Engl J Med (2013) 369:1691–1703. doi: 10.1056/NEJMoa1304369

21. Sherman RE, Anderson SA, Dal Pan GJ, Gray GW, Gross T, Hunter NL, et al. Real-world evidence — what is it and what can it tell us? N Engl J Med (2016) 375:2293–2297. doi: 10.1056/NEJMsb1609216

22. Gray WK, Navaratnam AV, Day J, Wendon J, Briggs TWR. Changes in COVID-19 in-hospital mortality in hospitalised adults in England over the first seven months of the pandemic: an observational study using administrative data. Lancet Reg Health Eur (2021) 5:100104. doi: 10.1016/j.lanepe.2021.100104

23. Kwok CS, Satchithananda D, Ahmed FZ, Chue CD, Barker D, Patwala A, et al. A critical evaluation of patient pathways and missed opportunities in treatment for heart failure. J Cardiovasc Dev Dis (2022) 9:455. doi: 10.3390/jcdd9120455

24. Schneeweiss S, Rassen JA, Brown JS, Rothman KJ, Happe L, Arlett P, et al. Graphical depiction of longitudinal study designs in health care databases. Ann Intern Med (2019) 170:398–406. doi: 10.7326/M18-3079

25. R Core Team. R: a language and environment for statistical computing. Vienna: R Foundation for Statistical Computing (2023). Available at: https://www.R-project.org/.

26. Partyka O, Pajewska M, Kwaśniewska D, Czerw A, Deptała A, Budzik M, et al. Overview of pancreatic cancer epidemiology in Europe and recommendations for screening in high-risk populations. Cancers (2023) 15:3634. doi: 10.3390/cancers15143634

27. Carrato A, Falcone A, Ducreux M, Valle JW, Parnaby A, Djazouli K, et al. A systematic review of the burden of pancreatic cancer in Europe: real-world impact on survival, quality of life and costs. J Gastrointest Cancer (2015) 46:201–211. doi: 10.1007/s12029-015-9724-1

28. King G, Ittershagen S, He L, Shen Y, Li F, Villacorta R. Treatment patterns in US patients receiving first-line and second-line therapy for metastatic pancreatic ductal adenocarcinoma in the real world. Adv Ther (2022) 39:5433–5452. doi: 10.1007/s12325-022-02317-9

29. Mackay TM, Latenstein AEJ, Bonsing BA, Bruno MJ, van Eijck CHJ, Koerkamp BG, et al. Nationwide compliance with a multidisciplinary guideline on pancreatic cancer during 6-year follow-up. Pancreatology (2020) 20:1723–1731. doi: 10.1016/j.pan.2020.10.032

30. Rahman RA, Moriarty M, McVey G, Ridgway P, Maguire D, Hoti E, et al. Outcome following centralisation of pancreatic cancer care in Ireland. Ann Oncol (2018) 29(Suppl 8):viii253. doi: 10.1093/annonc/mdy282.125

31. Gamboa AC, Rupji M, Switchenko JM, Lee RM, Turgeon MK, Meyer BI, et al. Optimal timing and treatment strategy for pancreatic cancer. J Surg Oncol (2020) 122:457–468. doi: 10.1002/jso.25976

32. Raptis DA, Fessas C, Belasyse-Smith P, Kurzawinski TR. Clinical presentation and waiting time targets do not affect prognosis in patients with pancreatic cancer. Surgeon (2010) 8:239–246. doi: 10.1016/j.surge.2010.03.001

33. Apollos JR, Sami S, Prasanth MN, Jeyakumar J, McFadyen AK. Pre-diagnostic delays caused by gastrointestinal investigations do not affect outcomes in pancreatic cancer. Ann Med Surg (2018) 34:66–70. doi: 10.1016/j.amsu.2018.07.011

34. Stornello C, Archibugi L, Stigliano S, Vanella G, Graglia B, Capalbo C, et al. Diagnostic delay does not influence survival of pancreatic cancer patients. United European Gastroenterol J (2020) 8:81–90. doi: 10.1177/2050640619879004

35. Jooste V, Dejardin O, Bouvier V, Arveux P, Maynadie M, Launoy G, et al. Pancreatic cancer: wait times from presentation to treatment and survival in a population-based study. Int J Cancer (2016) 139:1073–1080. doi: 10.1002/ijc.30166

36. Polonski A, Izbicki JR, Uzunoglu FG. Centralization of pancreatic surgery in Europe. J Gastrointest Surg (2019) 23:2081–2092. doi: 10.1007/s11605-019-04215-y

37. Hansen MFC, Storkholm JH, Hansen CP. The results of pancreatic operations after the implementation of multidisciplinary team conference (MDT): a quality improvement study. Int J Surg (2020) 77:105–110. doi: 10.1016/j.ijsu.2020.03.045

