## Supplementary Material 1 for "Claims-based reconstruction of the pancreatic cancer care pathway links centralization of care to survival"

**Supplementary Material 1 - Supplementary tables and figures**

**Table S1** Variables related to the patient pathway

| **Event** | **Examinations/therapies included** |
|---|---|
| IM | Computed tomography |
| | Magnetic resonance imaging |
| | Endoscopic ultrasonography |
| | Ultrasonography |
| | Nuclear imaging |
| HP | Histopathological examination |
| MDT | Multidisciplinary team |
| CTX | Chemotherapy: administration of a drug from the ‘L’ group according to the ATC classification |
| RT | External beam radiation therapy |
| | Brachytherapy |
| SX | Left-sided pancreatectomy |
| | Pancreaticoduodenectomy |
| | Total pancreatectomy |
| FLT | First line therapy: the first occurrence of SX, RT, or CTX in the sequence of events |
| COC | Complex oncological center: healthcare provider offering centralized treatment according to the Czech Society for Oncology |
| Non-COC | Other healthcare provider |

Notes: All other **IM** and **HP** events, with the exception of those representing the *index date* were ignored. For **MDT**, **SX**, **RT** and **CTX** events, only the first occurrence of the same event after the *index date* was retained; subsequent occurrences were ignored.

Abbreviations*: IM* imaging; *ATC* Anatomical Therapeutic Chemical classification; *RT* radiotherapy; *SX* surgery.

**Table S2** Median survival of treated pancreatic cancer patients according to the patient pathway ($N$ = 920)

| Pathway | *N* | Median survival (95% CI), days |
|---|---|---|
| IM_HP → MDT → SX | 169 | 1333 (1037–nr) |
| IM_HP → SX | 166 | 1130 (856–nr) |
| IM_HP → CTX | 305 | 296 (276–358) |
| IM_HP → RT | 13 | 278 (225–nr) |
| IM_HP → MDT → CTX | 253 | 360 (322–419) |
| IM_HP → MDT → RT | 14 | 753 (525–nr) |

Abbreviations: *CTX* chemotherapy; *HP* histopathological examination; *IM* imaging; *MDT* multidisciplinary team; *nr* not reached; *RT* radiotherapy;
*SX* surgery.

**Table S3** Adjusted survival of patients with pancreatic cancer according to the treatment strategy

| **Characteristic** | **SX model** | | **CTX model** | |
|---|---|---|---|---|
| | **HR (95% CI)** | ***P*-value** | **HR (95% CI)** | ***P*-value** |
| MDT review | 0.95 (0.67–1.34) | 0.8 | 0.88 (0.73–1.07) | 0.2 |
| Treated in COC | 0.68 (0.47–0.99) | 0.043 | 0.78 (0.63–0.97) | 0.023 |

Notes: Cox proportional hazards regression, adjusted by age.

Abbreviations*:* *CI* confidence interval; *COC* complex oncological center; *HR* hazard ratio; *MDT* multidisciplinary team.

**Table S4** Study limitations

| HAC data | Available | Not available | Available, not analyzed | Not available, classified due to study protocol |
|---|---|---|---|---|
| Sex | x | | | |
| Age | x | | | |
| Results of HP | | x | | |
| Presence of MDT review | x | | | |
| Disease stage | | x | | |
| Regimen of CTX | | | x | |
| Type of CTX and/or RT (neoadjuvant/adjuvant/ palliative) | | | | x |
| Type of SX | x | | | |
| Radicality of SX | | x | | |
| Centralization of care | x | | | |
| PS, comorbidities, patient preferences | | x | | |

Abbreviations*:* *CTX* chemotherapy; *HP* histopathological examination; *IM* imaging; *MDT* multidisciplinary team; *PS* performance status; *RT* radiotherapy; *SX* surgery.

**Figure S1** Time trends in MDT reporting for treated pancreatic cancer patients (*N* = 920)

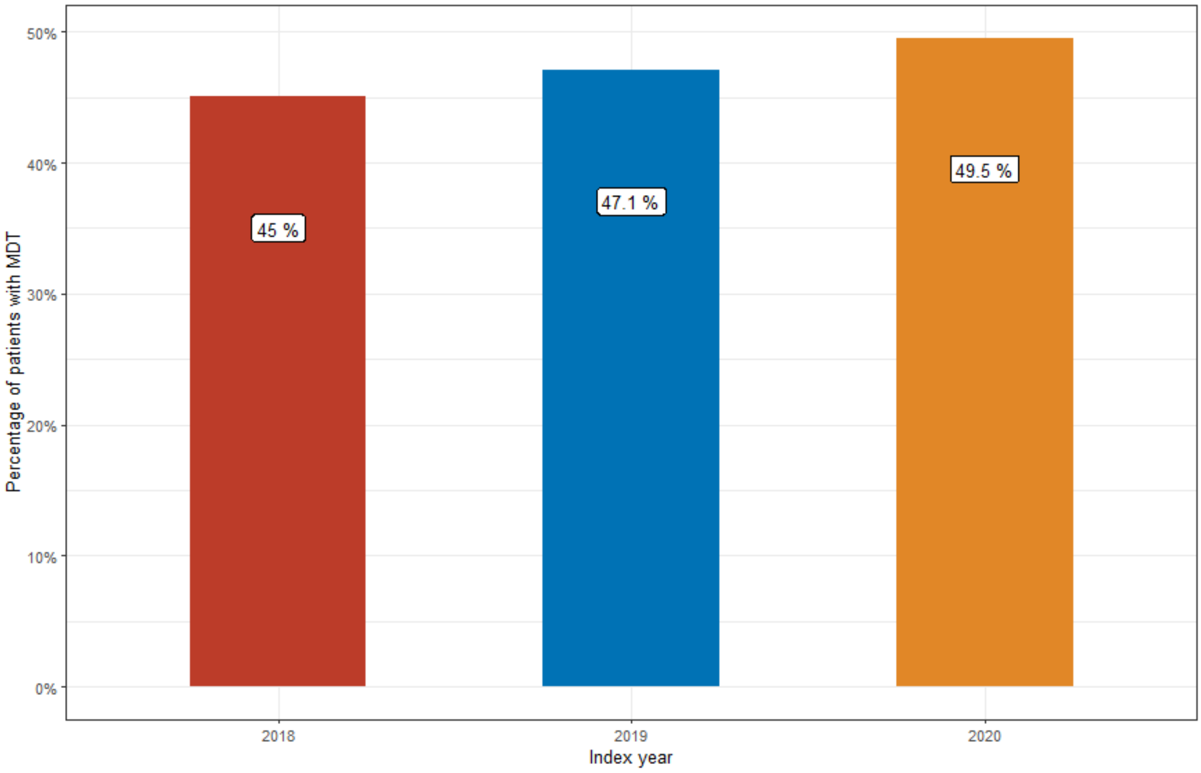


Notes: MDT was performed in 47.4% of patients, with this number rising from 45% in 2018 to 49% in 2020 for treated patients.

Abbreviations: *MDT* multidisciplinary team.

**Figure S2** Survival curves of treated pancreatic cancer patients according to the patient pathway (*N* = 920)

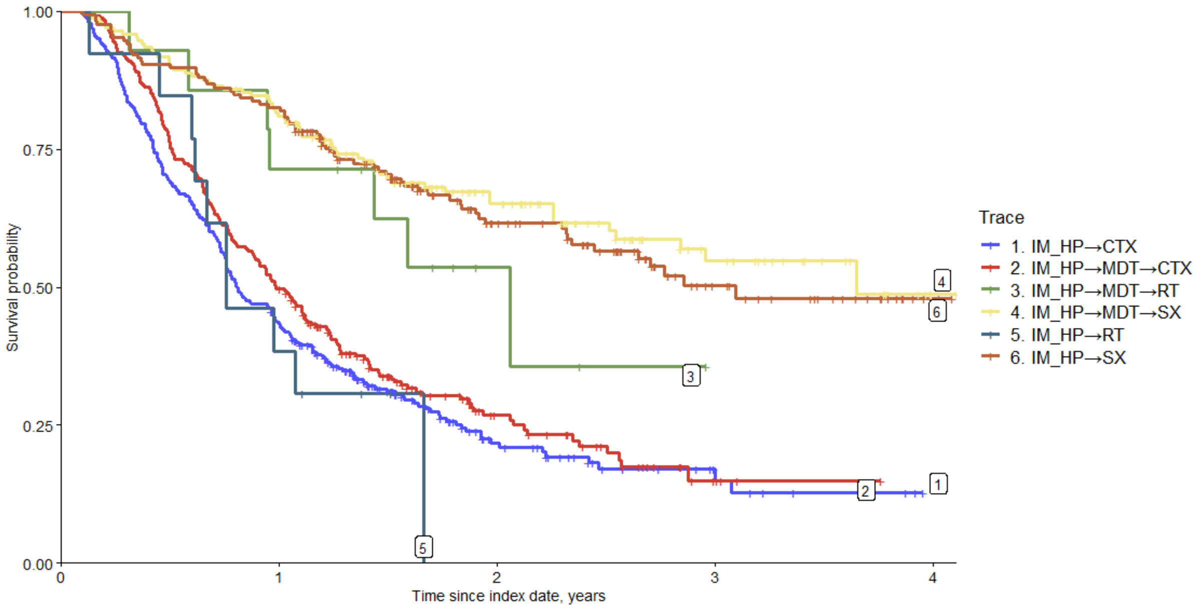


Abbreviations*:* *CTX* chemotherapy; *HP* histopathological examination; *IM* imaging; *MDT* multidisciplinary team; *RT* radiotherapy; *SX* surgery.

**Figure S3** Survival curves of treated pancreatic cancer patients according to centralization of care and the type of therapy

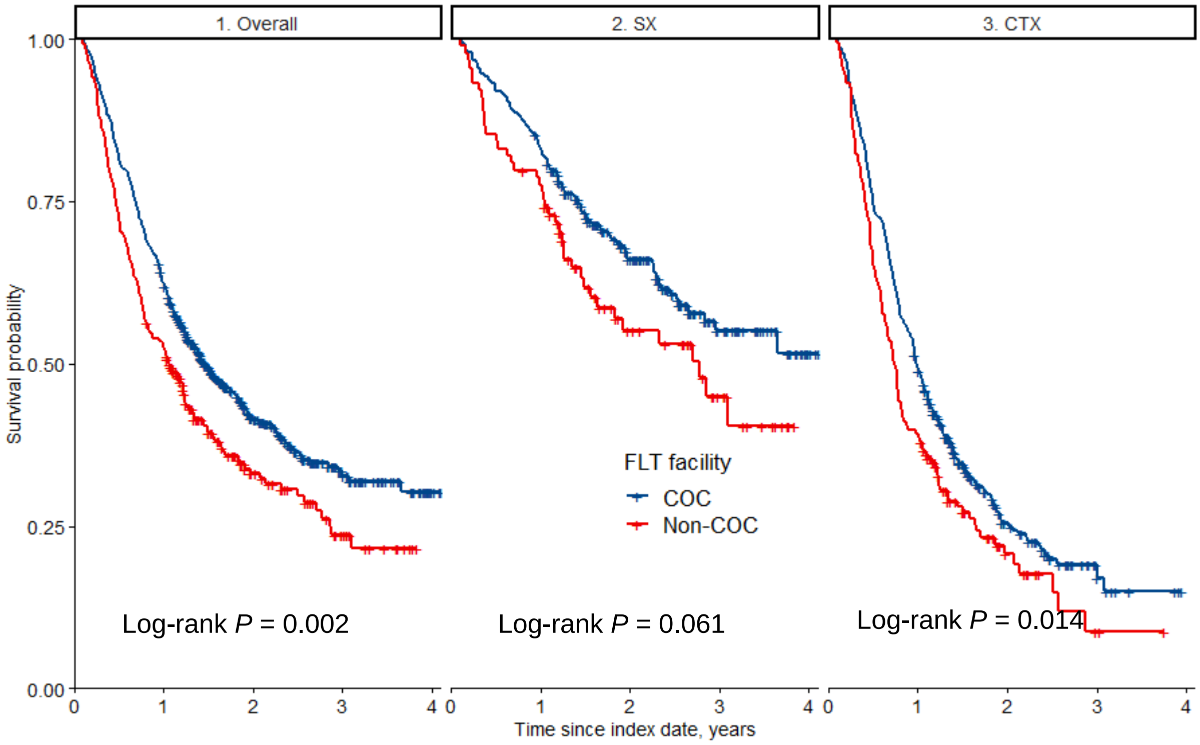


Abbreviations*:* *COC* complex oncological center*; CTX* chemotherapy; *FLT* first line therapy*; SX* surgery.

**Figure S4** Unadjusted survival of treated pancreatic cancer patients. (a) Effect of MDT on overall survival. (b) Effect of centralization of care on overall survival

(a) (b)

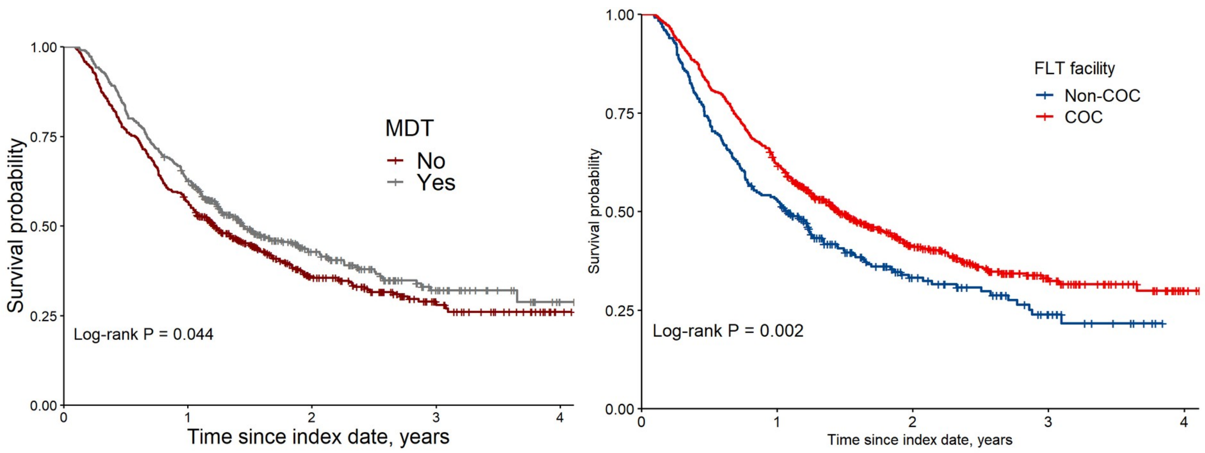


Abbreviations*:* *COC* complex oncological center; *FLT* first line therapy; *MDT* multidisciplinary team.
