## Supplementary Material 2 for "Claims-based reconstruction of the pancreatic cancer care pathway links centralization of care to survival"

### Supplementary material 2 - STROBE Statement checklist

Checklist of items that should be included in reports of cohort studies, completed for: “Claims-based reconstruction of the pancreatic cancer care pathway links centralization of care to survival”. Item wording is summarised; the full STROBE statement is available at https://www.strobe-statement.org.

| Item | Topic | Recommendation | Reported in |
|---|---|---|---|
| **Title and abstract** | | | |
| 1 | Title and abstract | Study design indicated in the title and abstract; structured summary of design, methods, results | Abstract, p. 1 |
| **Introduction** | | | |
| 2 | Background/rationale | Scientific background and rationale for the investigation | Introduction, pp. 1-2, paragraphs 1-7 |
| 3 | Objectives | Specific objectives and any prespecified hypotheses | Introduction, final paragraph p. 2; Methods, Study outcomes, pp. 3-4 |
| **Methods** | | | |
| 4 | Study design | Key elements of study design presented early in the paper | Study design and data source, p. 2. |
| 5 | Setting | Setting, locations, and relevant dates of recruitment, exposure, follow-up, data collection | Methods, Study design and data source, p. 2; Patient selection, p. 3; Figure 1, p. 13 |
| 6 | Participants | Eligibility criteria, sources and methods of selection of participants; methods of follow-up | Methods, Patient selection, p. 3; Figures 1-2, pp. 13-14 |
| 7 | Variables | Outcomes, exposures, predictors, potential confounders, effect modifiers defined | Methods, Study outcomes, pp. 3-4; Table S1 (Supplementary Material 1, pp. 1-2) |
| 8 | Data sources / measurement | Sources of data and details of methods of assessment for each variable | Methods, Study design and data source, p. 2; Table S1 (Supplementary material 1, pp. 1-2) |
| 9 | Bias | Efforts to address potential sources of bias | Methods, Patient selection (misclassification, washout), p. 3; Discussion, Limitations, p. 7 |
| 10 | Study size | How the study size was arrived at | Methods, Patient selection, p. 3; Results, Patients, p. 4; Figure 2, p. 14 |
| 11 | Quantitative variables | Handling of quantitative variables and grouping used | Methods, Statistical analysis, p. 4 |

| Item | Topic | Recommendation | Reported in |
|---|---|---|---|
| 12 | Statistical methods | All statistical methods, including those used to control for confounding; subgroups and interactions; missing data; loss to follow-up; sensitivity analyses | Methods, Statistical analysis, p. 4 |
| **Results** | | | |
| 13 | Participants | Numbers at each stage; reasons for non-participation; flow diagram | Results, Patients, p. 4; Figure 2-3, p. 14 |
| 14 | Descriptive data | Characteristics of study participants; missing data for each variable; follow-up time | Results, Patients, p. 4; Methods, Study outcomes, pp. 3-4 |
| 15 | Outcome data | Numbers of outcome events or summary measures over time | Results, Patient pathways, p. 4; Patient prognosis, p. 5; Figure 3-4, pp. 14-15; Table 1, p. 12 |
| 16 | Main results | Unadjusted and confounder-adjusted estimates with precision; confounders adjusted for; category boundaries | Results, Patient prognosis, p. 5; Table 1, p. 12; Table S3 and Figure S4 (Supplementary Material 1, pp. 4 and 9, respectively) |
| 17 | Other analyses | Other analyses performed –subgroups, interactions, sensitivity analyses | Results, Time to treatment, p. 5; Figure S1-S3 (Supplementary Material 1, pp. 6-8) |
| **Discussion** | | | |
| 18 | Key results | Key results summarised with reference to study objectives | Discussion, paragraphs 1-2, p. 5; Conclusion, p. 8 |
| 19 | Limitations | Limitations, sources of potential bias or imprecision, direction and magnitude of bias | Discussion, Limitations of the study, p. 7 |
| 20 | Interpretation | Cautious overall interpretation considering objectives, limitations, multiplicity, and other evidence | Discussion, paragraphs 3-12, pp. 5-7; Conclusion, p. 8 |
| 21 | Generalisability | Generalisability (external validity) of study results | Discussion, paragraphs 2, 5, pp. 5-6; Limitations, p. 7 |
| **Other information** | | | |
| 22 | Funding | Source of funding and role of funders | Funding statement, p. 8 |
